# The Effect of Non-Pharmacologic Strategies on Prevention or Management of Intensive Care Unit Delirium: A Systematic Review

**DOI:** 10.1101/2020.05.20.20100552

**Authors:** Haleh Hashemighouchani, Julie Cupka, Jessica Lipori, Matthew M Ruppert, Tezcan Ozrazgat-Baslanti, Parisa Rashidi, Azra Bihorac

## Abstract

**Objective:** The aim of this systematic review is to examine and report on recently available literature evaluating the relationship between non-pharmacologic management strategies and the reduction of delirium in the intensive care unit.

**Methods:** Six major research databases were systematically searched for articles analyzing the efficacy of non-pharmacologic delirium interventions in the past five years. Search results were restricted to adult human patients aged 18 years or older in the intensive care unit setting, excluding terminally ill subjects and withdrawal-related delirium. Following title, abstract, and full text review, 27 articles fulfilled the inclusion criteria and are included in this report.

**Results:** The 27 reviewed articles consist of 12 interventions with a single-component investigational approach, and 15 with multi-component bundled protocols. Delirium incidence was the most assessed outcome followed by duration. Family visitation was the most effective individual intervention and mobility interventions were the least effective. Two of the three family studies significantly reduced delirium incidence, while one in five mobility studies did the same. Multi-component bundle approaches were the most effective of all; of the reviewed studies, eight of 11 bundles significantly improved delirium incidence and seven of eight bundles decreased the duration of delirium.

**Conclusions:** Multi-component, bundled interventions were more effective at managing intensive care unit delirium than those utilizing an approach with a single interventional element. Although better management of this condition suggests a decrease in resource burden and improvement in patient outcomes, comparative research should be performed to identify the importance of specific bundle elements.

## 1. Introduction

Delirium is a multifactorial, acute, confusional state characterized by the disturbance of consciousness and cognition; it is particularly common in the intensive care unit (ICU) with incidence ranging from 19 to 87% with especially high rates in mechanically ventilated patients [1-3]. ICU delirium is associated with adverse outcomes including increased mortality, prolonged mechanical ventilation and hospitalization, increased risk of cognitive dysfunction after discharge, and increased cost of care [4-6].

While the pathophysiology of delirium is not well understood, there are multiple factors associated with an increased risk for developing delirium including age, neurologic or psychological disorders, polypharmacy, medications, and sensory impairment [7-9]. Modifiable environmental risk factors including immobilization, use of restraints, isolation, and levels of environmental light and sound are also considered risk factors for the development of delirium in the ICU [7, 10].

The morbidity associated with delirium as well as the multitude of delirium risk factors present in the ICU make delirium prevention and management strategies essential. These strategies have included pharmacological, non-pharmacological, and multicomponent interventions with the aim of decreasing the incidence and duration of delirium. Research into pharmacological interventions has focused on haloperidol and dexmedetomidine, though there has also been limited research into the effects of ramelteon, melatonin, and ziprasidone [11-14]. Despite continued research, current literature does not support the use of anti-psychotic agents, benzodiazepines, or melatonin in the management of delirium [11, 15].

Given the lack of evidence supporting pharmacological measures, further research into the efficacy of non-pharmacologic interventions such as early mobilization, environmental modifications, or management bundles is crucial. Implementing effective delirium management shows promise in decreasing morbidity, mortality, length of stay, and resource burden in the ICU setting. In terms of the PICOS framework (Population, Interventions, Comparisons, Outcomes, Study Design) [56], our systematic review aims to address the effects of any non-pharmacologic prevention or management strategy on the incidence, prevalence, duration, or severity of delirium in critically ill adult patients compared to control patients, with no restrictions on study design.

## 2. Materials and Methods

### 2.1 Search strategy and Data Extraction

PRISMA guidelines were followed in this review and attached as Supplementary Appendix 1 [16]. The electronic databases of PubMed, Embase, Cochrane Central, Web of Science, CINAHL, and ClinicalTrials.gov were systematically searched for articles concerning non-pharmacologic treatments for delirium in the ICU. Search terms were tailored to each database in order to best utilize the individual subject headings, keywords, and MeSH terms included in the individual databases. A full list of search terms is shown in Supplementary Appendix 2.

In addition to our search terms, search results were restricted to articles published in English within the past five years (Jan 1, 2014 to May 15, 2019). After search results were compiled and duplicates were removed, a total of 5234 articles were selected for title and abstract review. Two pairs of independent authors screened the titles and abstracts and retrieved articles for eligibility resulting in 113 articles selected for full text review. The same two authors independently reviewed the full text of eligible articles, completed data extraction worksheets adapted from the Cochrane Review Group’s Data Extraction Form [17], and assessed the articles for risk of bias using the Cochrane risk of bias tool [18]. Elements of the data extraction worksheet included study design and setting, participant characteristics, details of the intervention and control groups, diagnostic tools, and patient outcomes (Supplementary Table 1). Any disagreements were resolved by thoroughly discussing any points of concern. During full text review 86 articles were removed because they failed to meet our inclusion criteria resulting in a total of 27 included articles (Figure 1).

**Figure 1.**
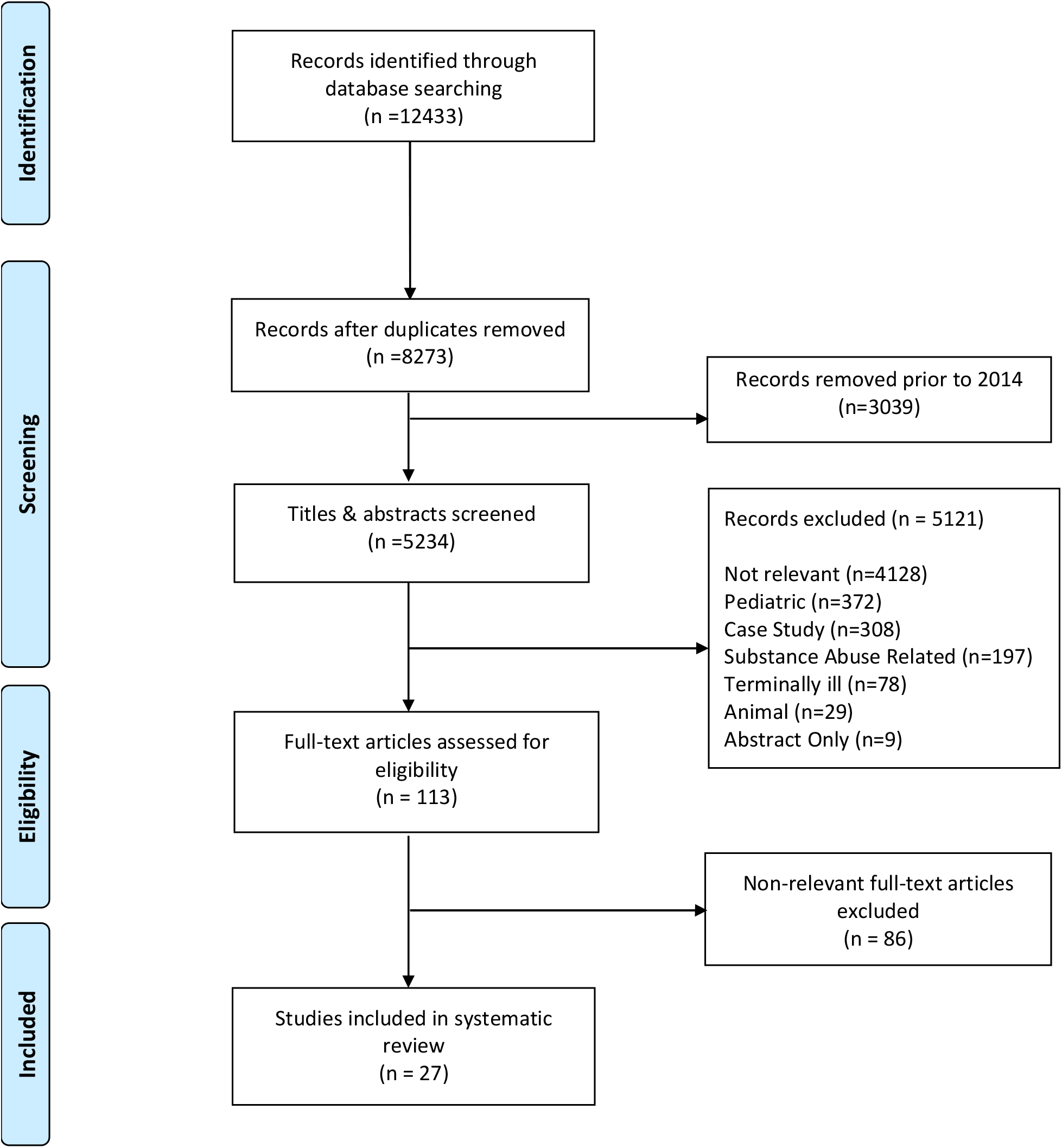
PRISMA Record Screening Flow Chart

**Table 1.**
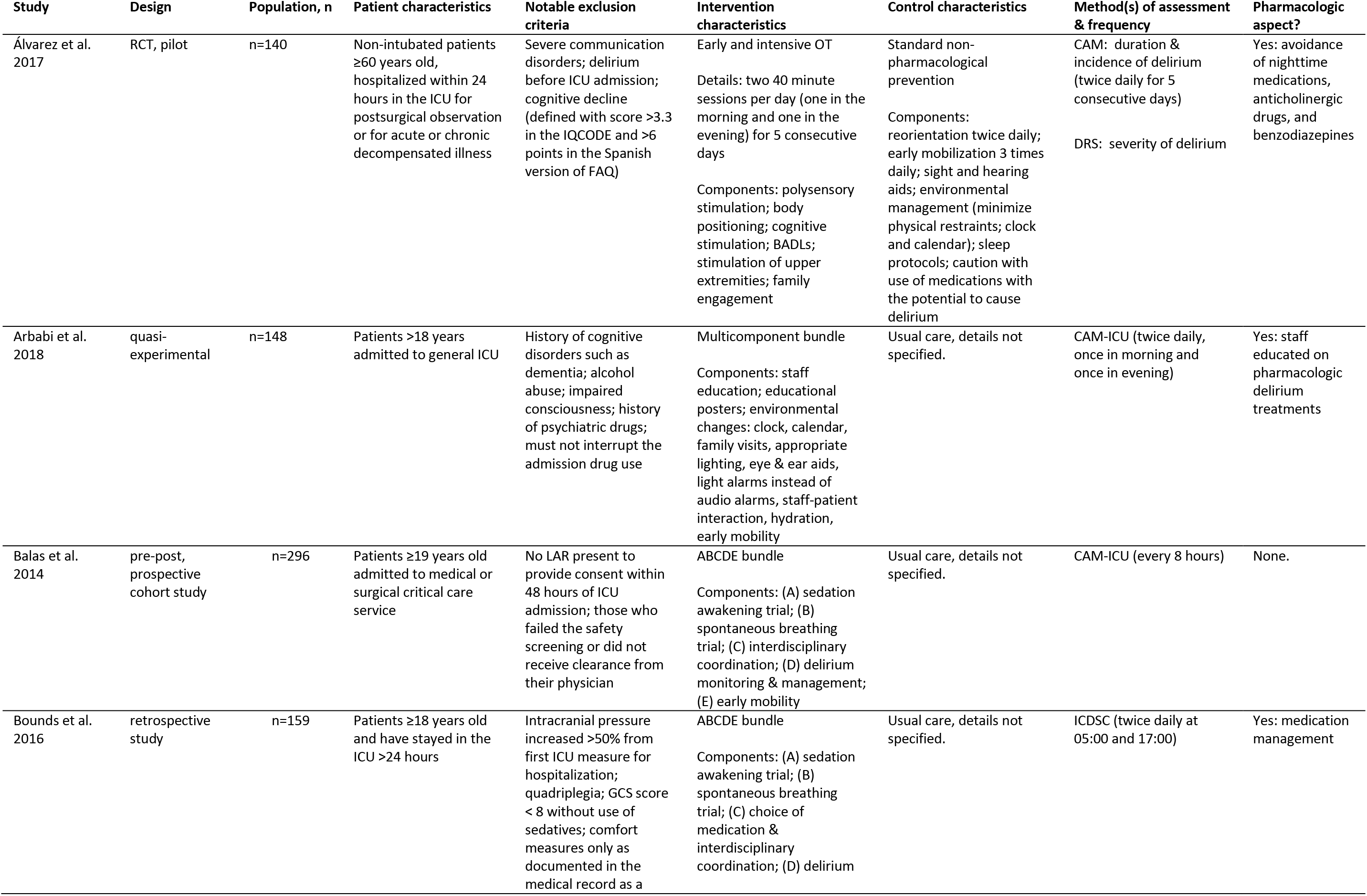
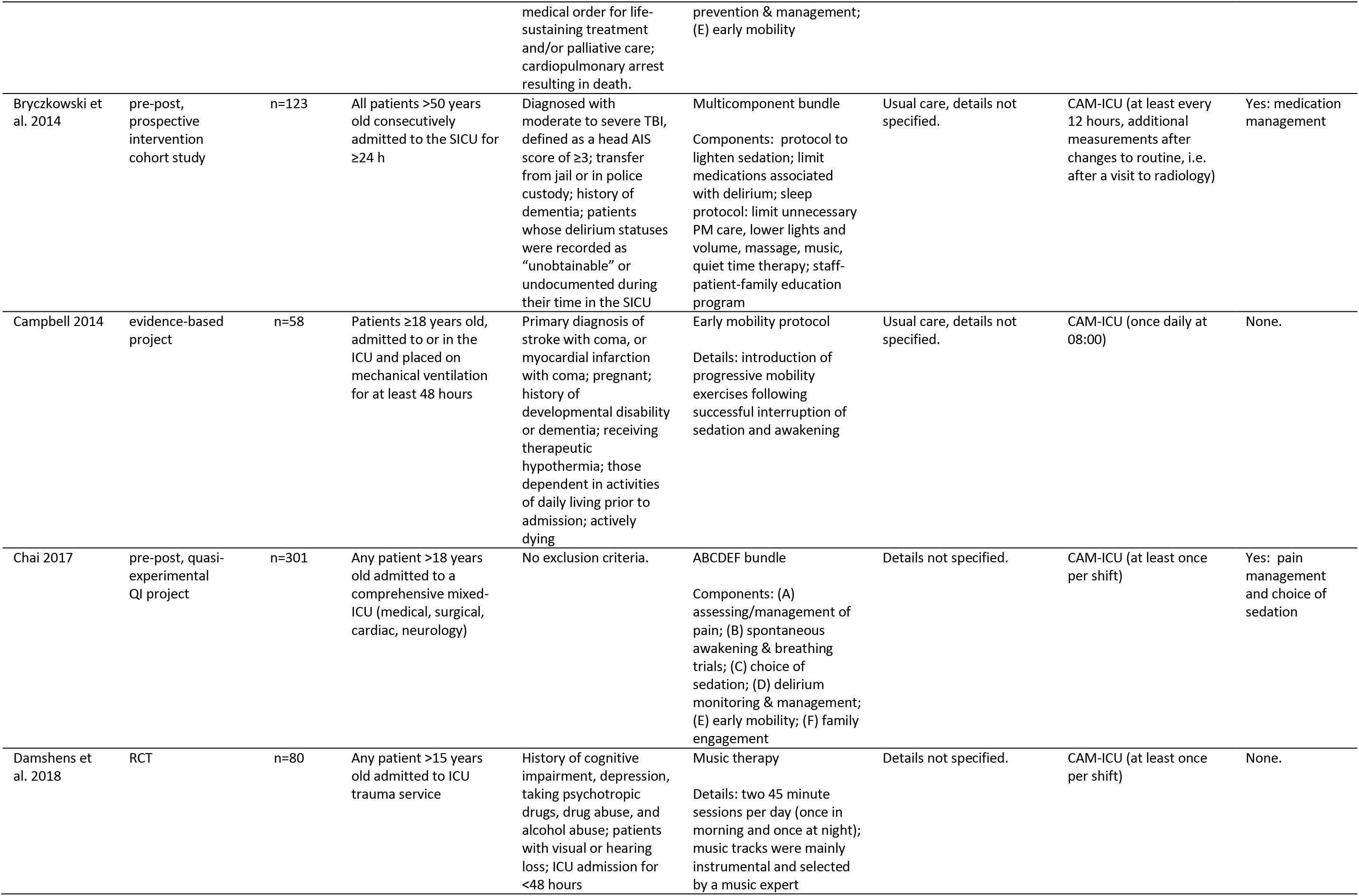
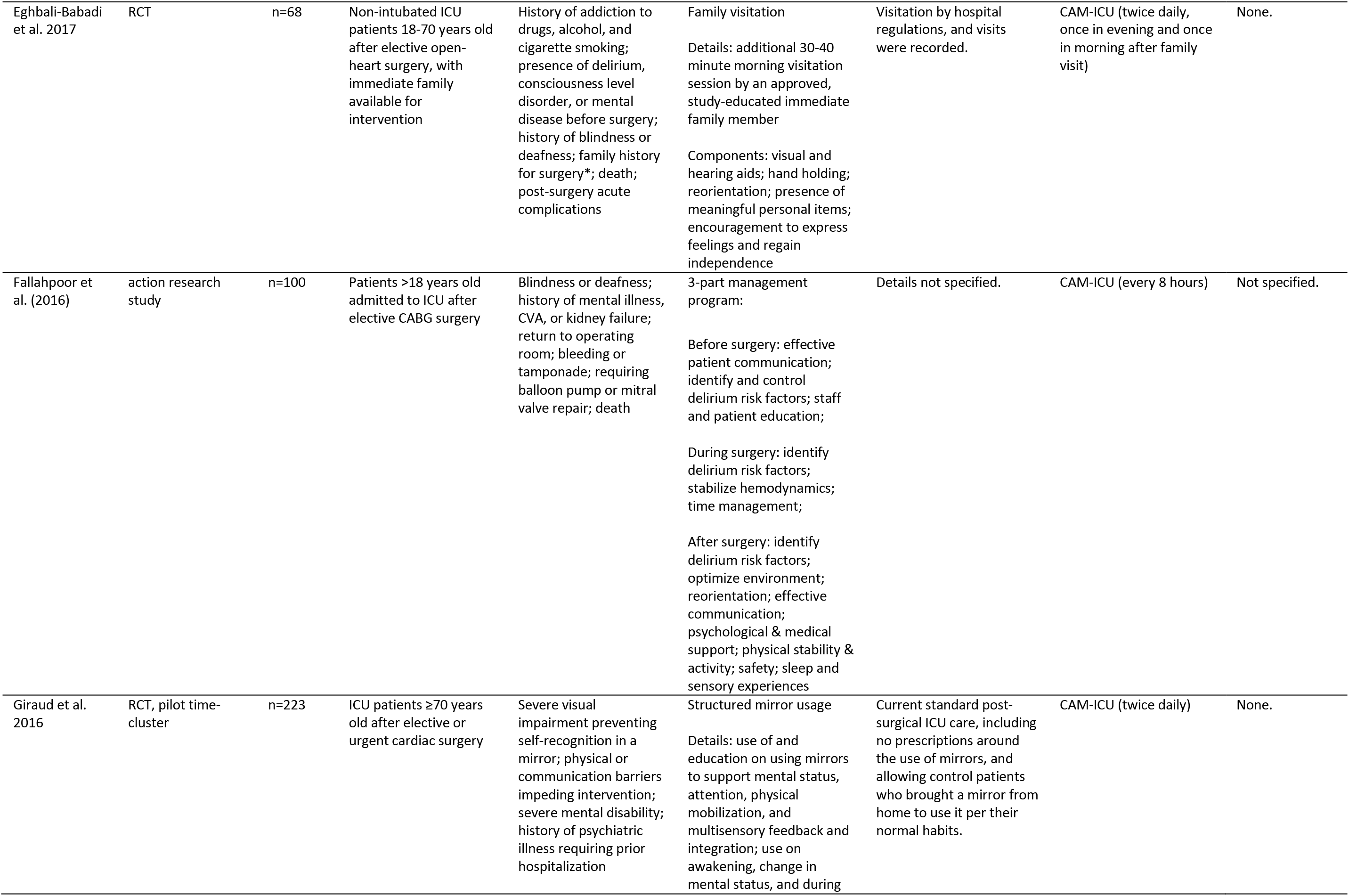
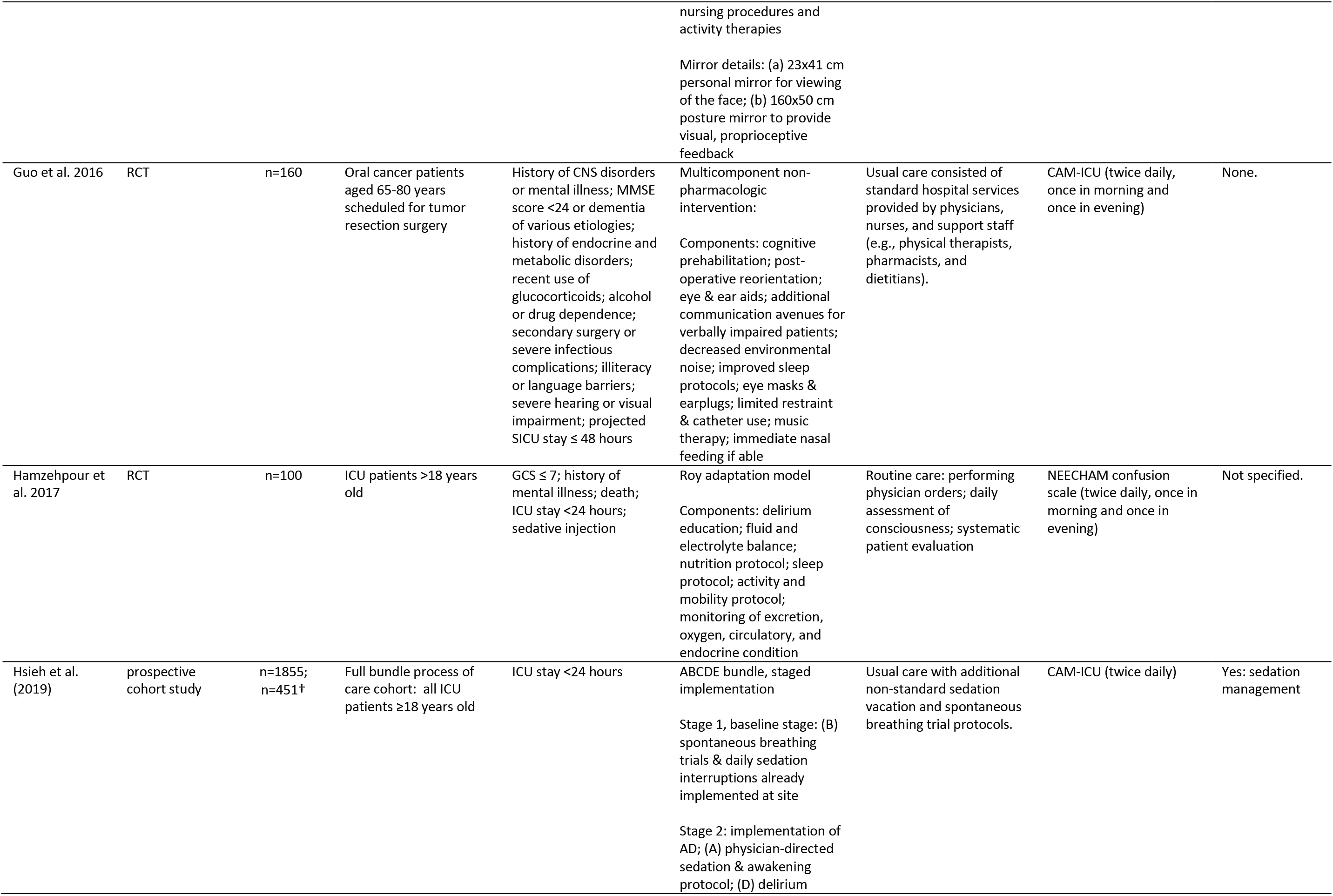
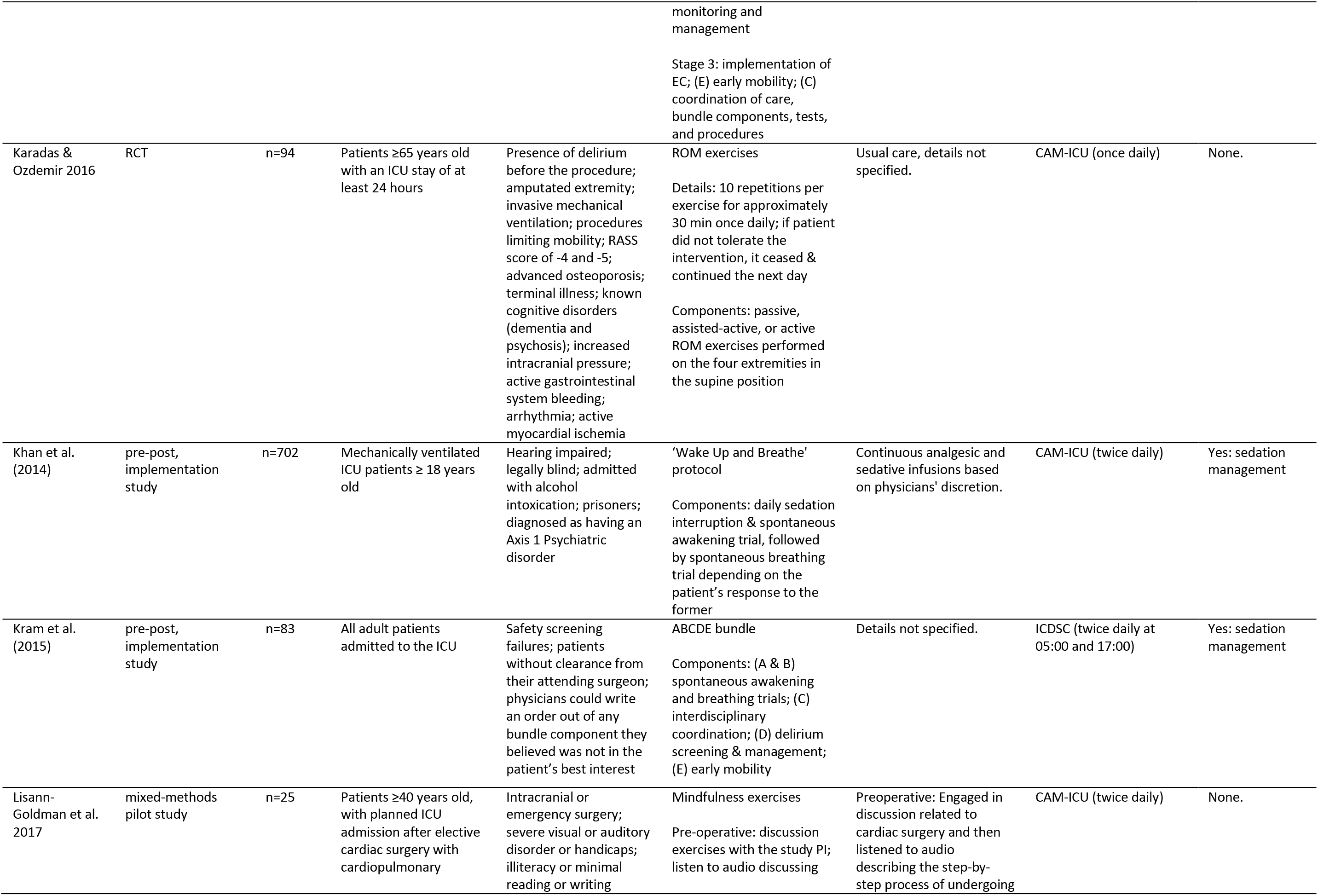
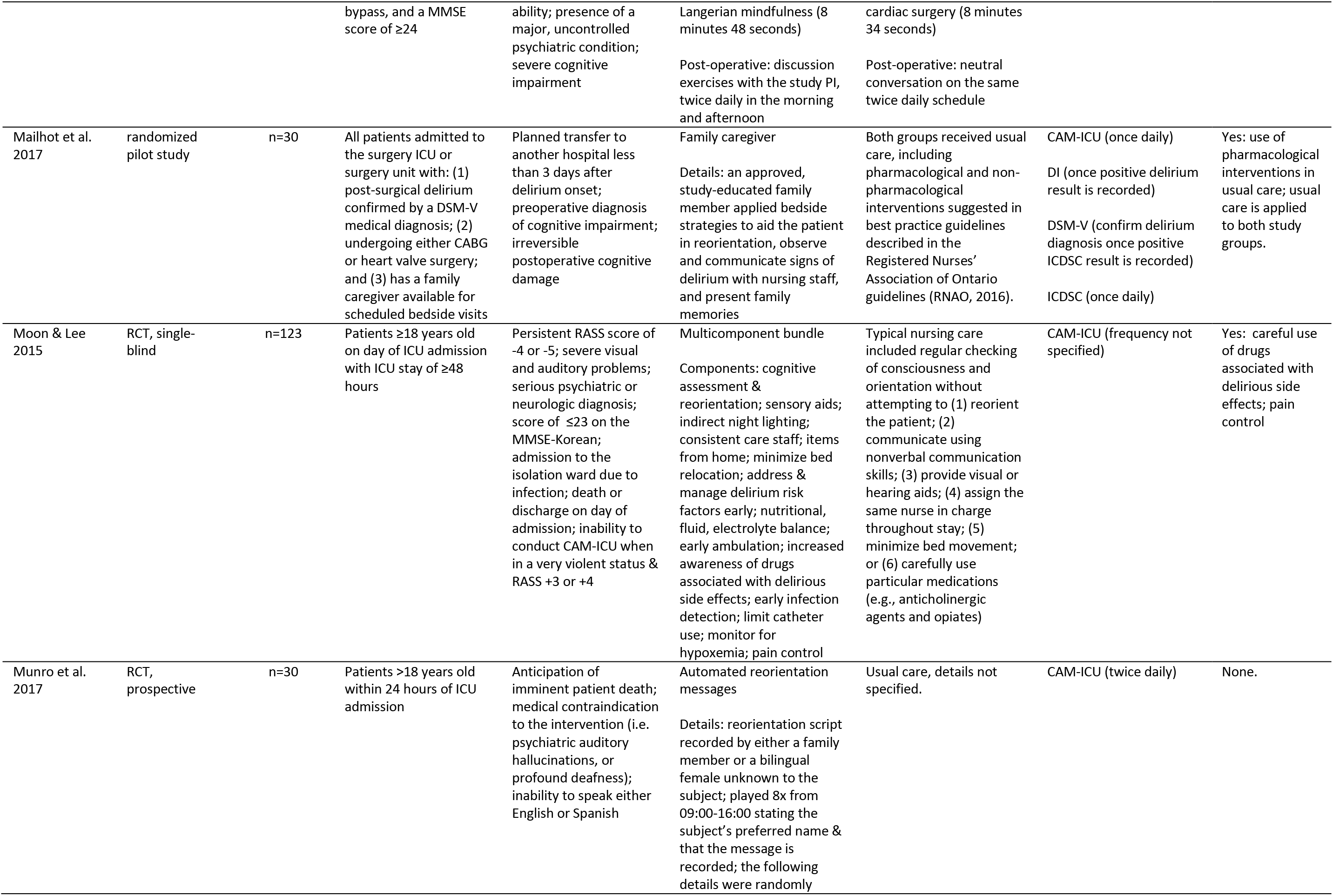
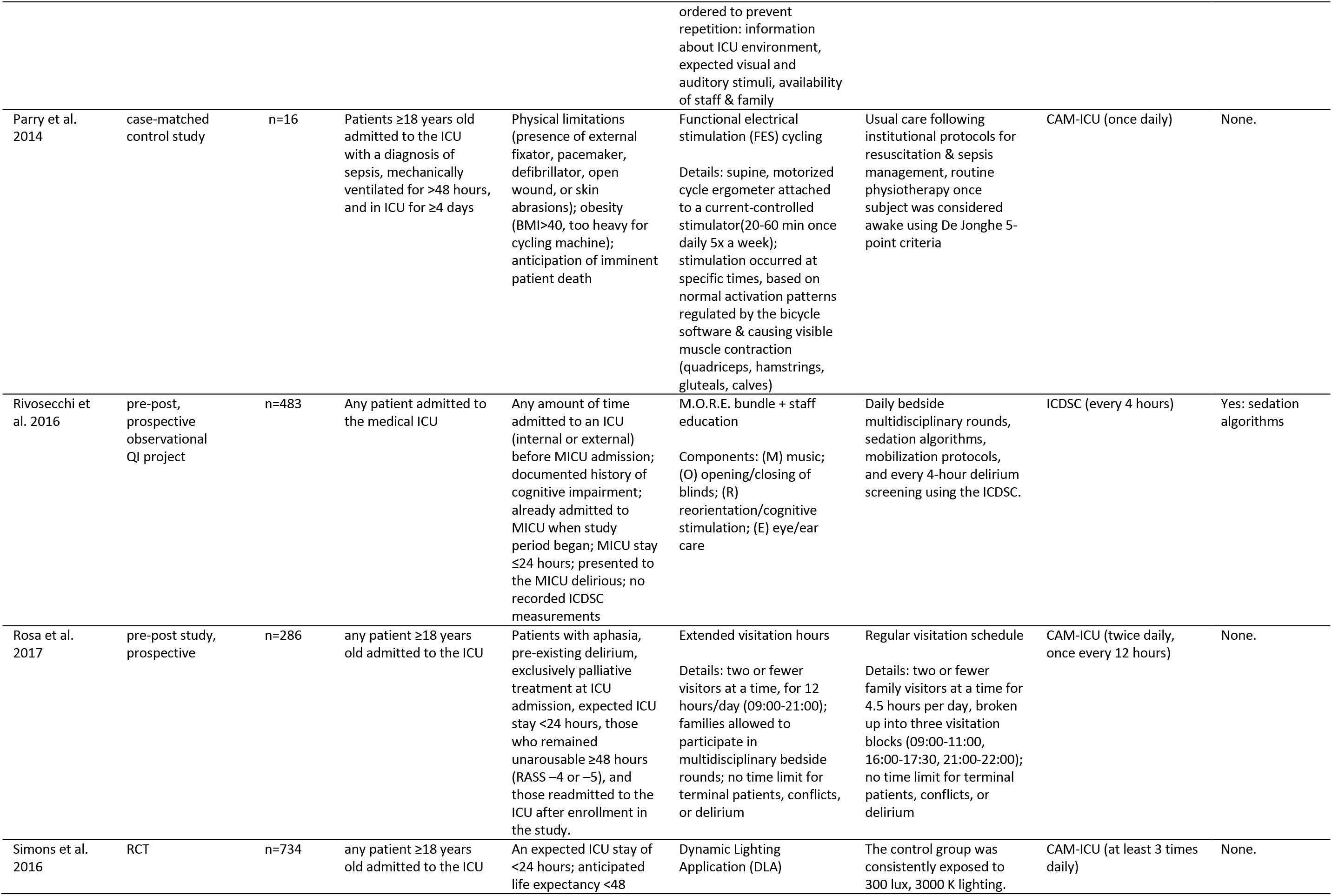
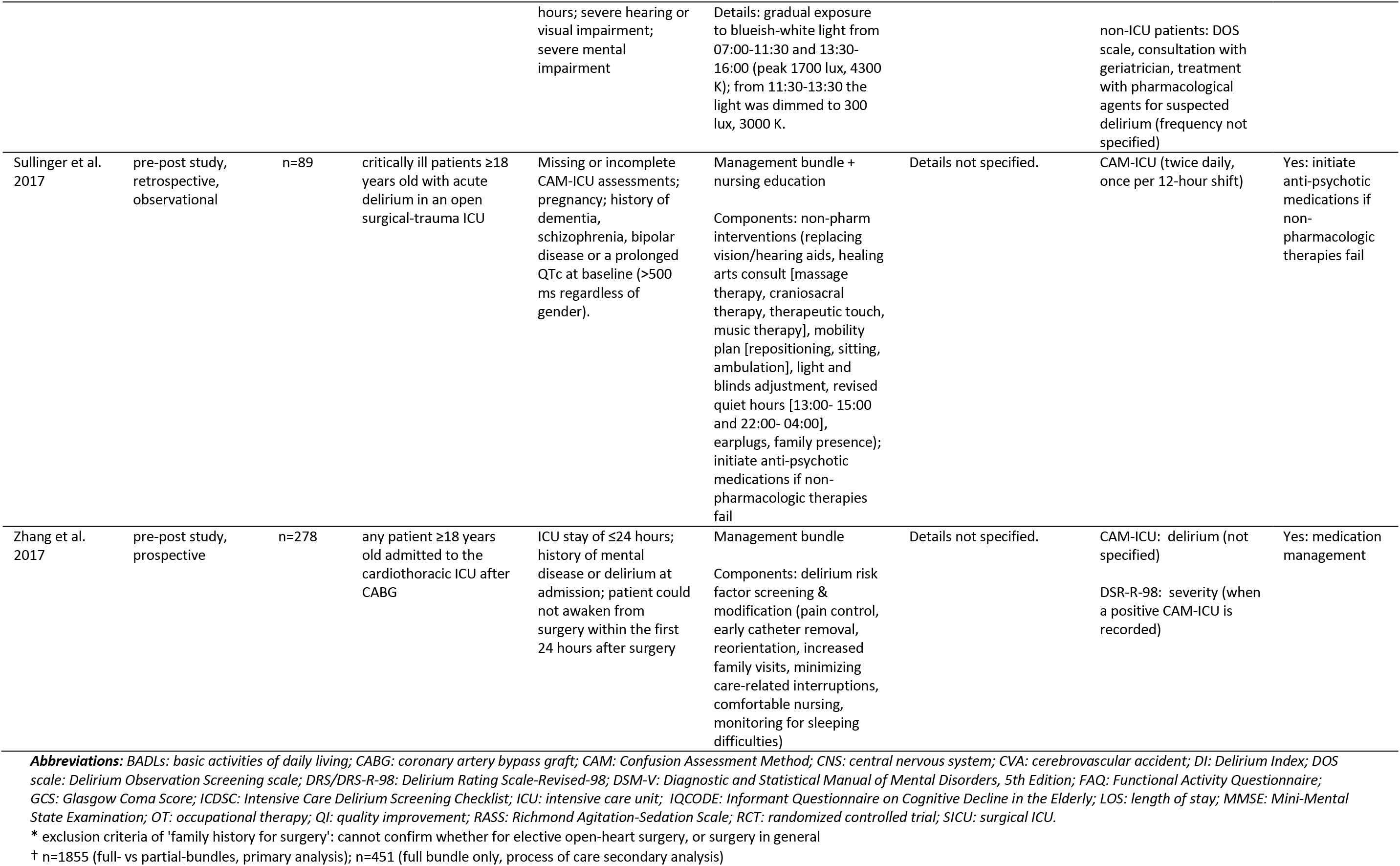
Summary of Study Characteristics

### 2.2 Inclusion and Exclusion Criteria

Our review addresses non-pharmacological management strategies for delirium in the ICU. Included articles were those investigating non-pharmacologic interventions and their impact on delirium incidence, prevalence, duration, or severity in an adult (≥ 18 years) intensive care unit setting. Articles were excluded if they focused on non-human subjects, pediatrics, terminally ill subjects, withdrawal related delirium, case reports, or where no full-text article was available (abstract only). There were no restrictions on study design. Studies solely investigating delirium-free-coma-free days were excluded since it is not possible to review as a delirium-specific result. One multi-center study was excluded as both the frequency and method of assessment for delirium was not specified for all study centers, making it difficult to reliably compare the results with other trials [19]. Another study was excluded because neither the screening process nor the cohort were described other than total number of patients enrolled, and there were no exclusion criteria noted to infer any characteristics of the selected population [20].

### 2.3 Risk of Bias Assessment

In addition to data extraction using the Cochrane Review Group’s Data Extraction Form [17], a risk of bias assessment was performed by all authors on all included RCT and randomized pilot studies. A risk of bias worksheet was developed by modifying Cochrane’s Risk of Bias Tool and articles were ranked as having a low, high, or unclear risk of bias [18]. Disagreements were settled by discussion between the authors. A total of eleven included studies underwent this assessment. Details of the risk of bias assessment categories can be found in Supplementary Table 2.

**Table 2.** Summary of Delirium Outcomes

| **Individual Interventions** | **Incidence** | **Prevalence** | **Duration** | **Severity** |
| --- | --- | --- | --- | --- |
| automated reorientation [41] | -- | -- | y/x | -- |
| dynamic lighting [45] | x | -- | x | -- |
| family, caregiver [39] | o | -- | o | x |
| family, RVM vs EVM [44] | y | -- | -- | -- |
| family, additional structured visit [29] | y | -- | -- | -- |
| mindfulness exercises [38] | o | -- | -- | -- |
| mirrors [31] | x | -- | x | -- |
| mobility, early and intensive OT [21] | y | -- | y | x |
| mobility, early [26] | x | -- | x | -- |
| mobility, ROM exercises [35] | x | -- | x | -- |
| mobility, FES [42] | x | -- | y | -- |
| music therapy [28] | x | -- | -- | -- |
| **Bundled Interventions** | **Incidence** | **Prevalence** | **Duration** | **Severity** |
| ABCDE [23] | -- | y | y/x | -- |
| ABCDE [24] | -- | y | y | -- |
| ABCDE [37] | -- | o | -- | -- |
| ABCDE 3 stage implementation [34] | y | -- | -- | -- |
| ABCDEF [27] | y | -- | -- | -- |
| M.O.R.E. [43] | y | -- | y | -- |
| multicomponent [22] | y | -- | y | -- |
| multicomponent [25] | x | -- | y | -- |
| multicomponent [40] | x | -- | -- | -- |
| multicomponent [46] | -- | -- | y | -- |
| multicomponent non-pharmacologic [32] | y/x | -- | y | -- |
| post-CABG delirium management [30] | y | -- | -- | -- |
| risk factor screening & target modification [47] | y | -- | x | x |
| Roy adaptation model [33] | y/x | -- | -- | y/x |
| 'Wake Up and Breathe' protocol [36] | x | x | -- | -- |

**Legend:** x: $p>0.05$ ; y: $p<0.05$ ; o: not analyzed for significance; --: not measured in this study; y/x: some measured time-points are significant.

## 3. Results

After searching the literature, 27 articles are included in our systematic review [21-47] (Figure 1). Study details of each reviewed trial are located in Table 1. The 27 included studies provide results on many distinct outcomes; however, only the delirium related outcomes of incidence, prevalence, duration, and severity were reviewed (Tables 2-5). Outcomes combining delirium and coma into the same statistic were excluded, as no delirium-specific results could be assessed outright.

Of these 27 articles, 24 assessed incidence and/or prevalence within their cohorts [21-40, 42-45, 47], 16 assessed for duration [21-25, 31, 32, 35, 39, 41-43, 45-47], and four for severity [21, 33, 39, 47]. Additionally, 12 focused on the effect of single interventions [21, 26, 28, 29, 31, 35, 38, 39, 41, 42, 44, 45] while 15 considered bundled, multicomponent interventions [22-25, 27, 30, 32-34, 36, 37, 40, 43, 46, 47]. Individual interventions included mobility protocols, distinct family visiting policies, dynamic lighting, music therapy, automated reorientation messages, mindfulness exercises, and the structured use of mirrors in recovery. These individual interventions also comprised multiple components of the bundled interventions.

Measurements for incidence, prevalence, and duration were based upon multiple methods of delirium screening, including the CAM, CAM-ICU, ICDSC, and NEECHAM scales. Incidence and prevalence were similarly defined in all studies except for one, but are recorded separately in Table 3; only one study looked at both incidence and prevalence [36]. Severity was assessed by using the Delirium Index (DI), the Delirium Rating Scale (DRS), the Revised Delirium Rating Scale (DRS-R-98), and NEECHAM scale (Table 5).

**Table 3.**
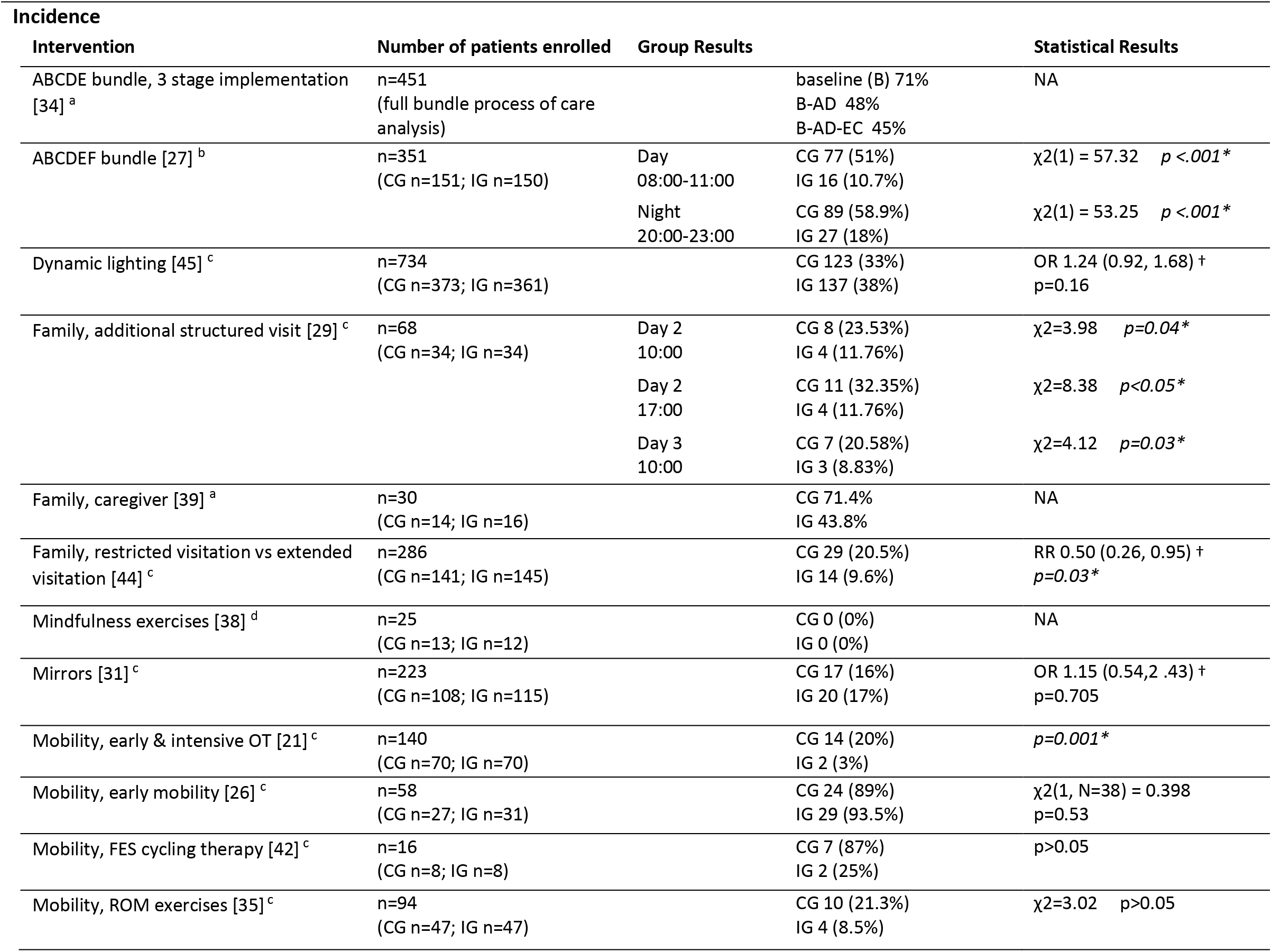
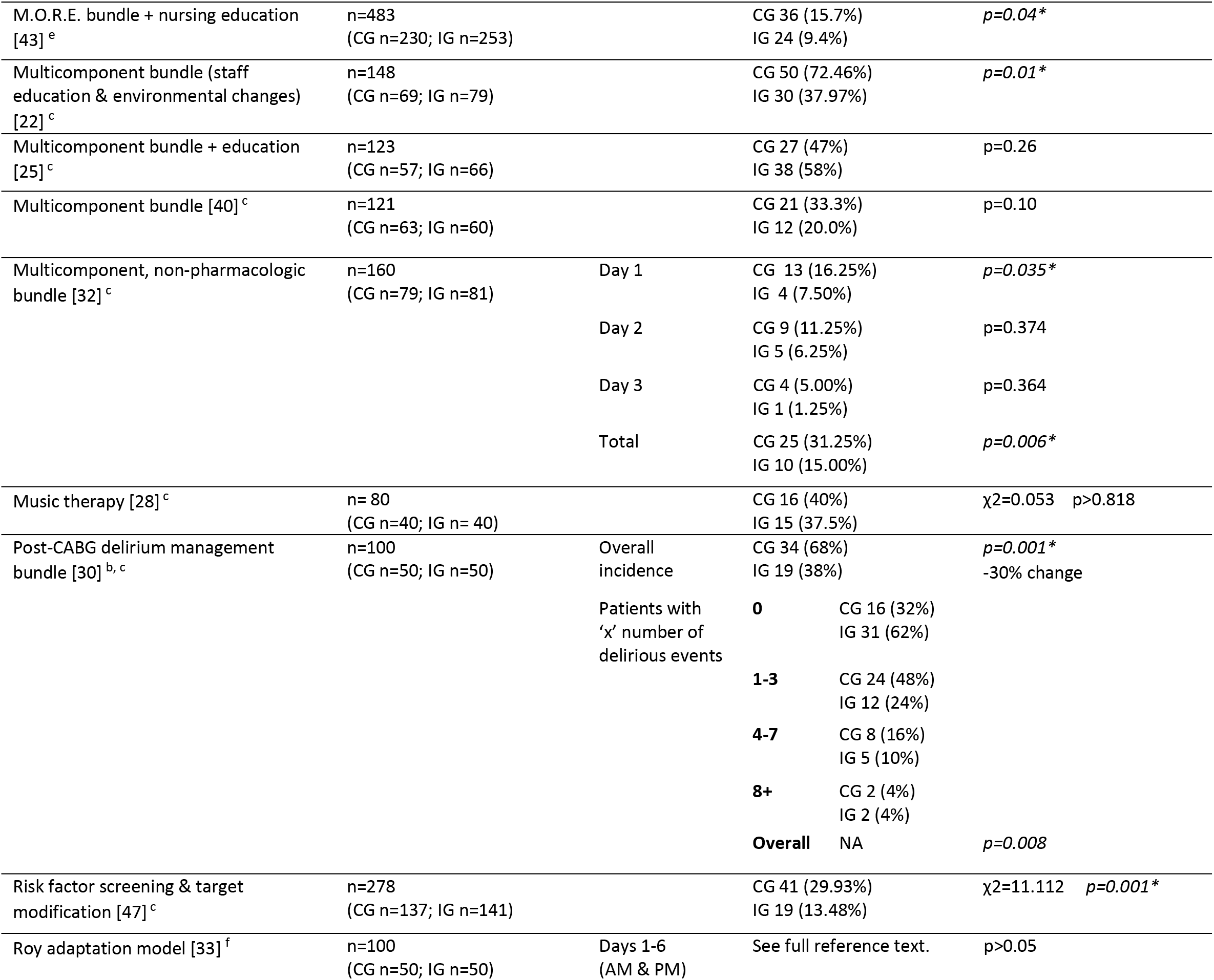
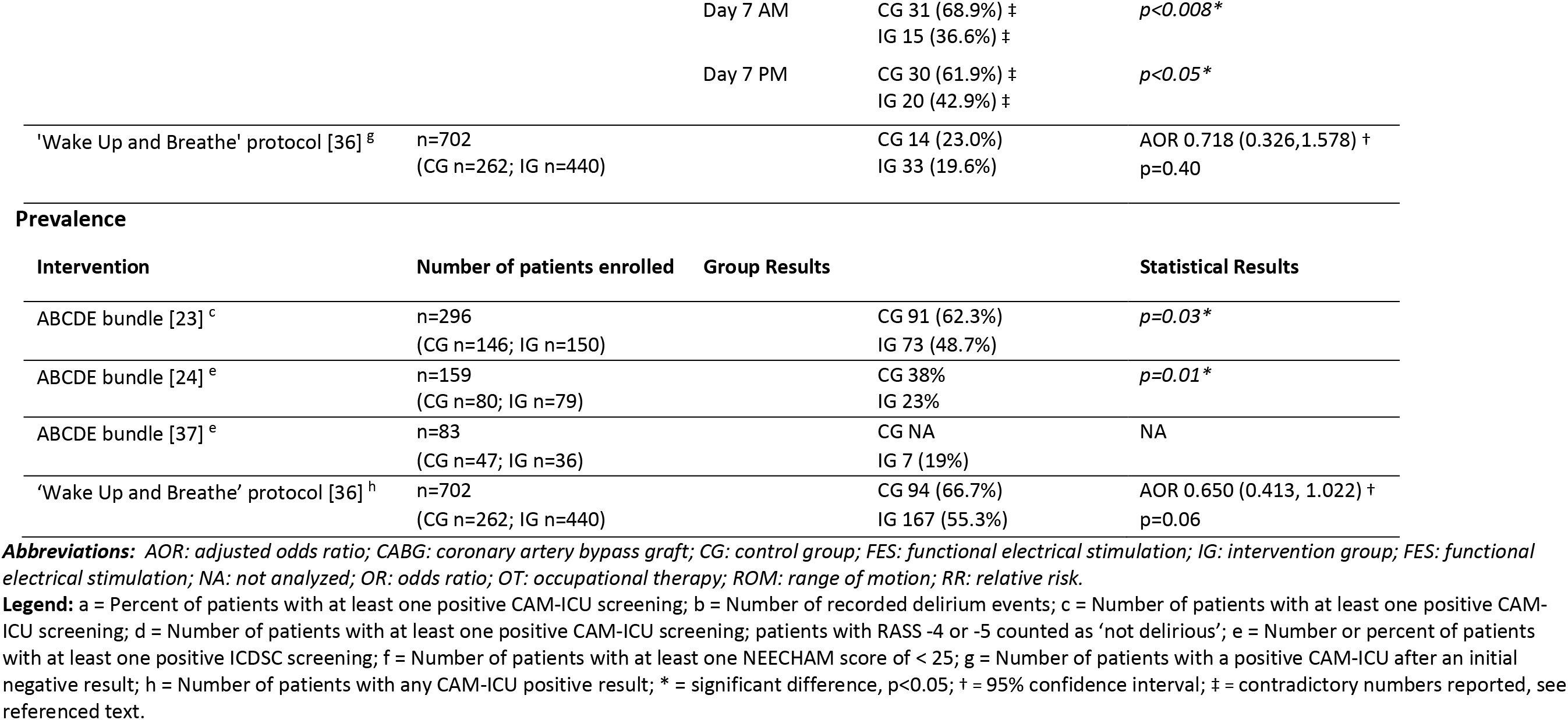
Results on Incidence and Prevalence of Delirium

Of the 27 included studies, 11 were RCTs or randomized pilot studies [21, 28, 29, 31-33, 35, 39-41, 45], eight were pre-post prospective studies [23, 25, 36, 37, 43, 44, 46, 47], two were quasi-experimental [22, 27], and the remaining six were a case-matched control study, an evidence based protocol, a mixed-methods pilot study, a prospective cohort study, a retrospective cohort study, and an action research study [24, 26, 30, 34, 38, 42].

### 3.1 Risk of Bias Assessment

The ten RCTs and the randomized pilot study underwent a risk of bias assessment performed by two independent authors (Supplementary Table 2). Risks of bias fell into five major groups (selection bias, performance bias, detection bias, attrition bias, and reporting bias), and based on a study’s scores in each of these groups it was labeled as having an overall high, low, or unclear risk of bias. Four were considered low risk of bias [21, 31, 39, 45], two had a high risk of bias [29, 41], and five had an unclear risk of bias [28, 32, 33, 35, 40]. The most common source of bias was performance bias due to the impossibility of blinding participants or personnel to certain treatments. Common sources of unclear and high risk of bias included the methods of randomization and allocation concealment, as well as how missing data was handled.

### 3.2 Individual interventions

#### 3.2.1 Early mobility

The effect of early mobility protocols on delirium was the most commonly studied individual intervention. Four of the studies included in our review individually assessed the efficacy of early mobility [21, 26, 35, 42]) in treating and preventing delirium; of these, two were RCTs [21, 35], one was an evidence-based project [26], and one was a case-matched control study [42]. They assessed delirium through CAM [21] and CAM-ICU [26, 35, 42].

The pilot RCT performed by Álvarez et al. investigated the effect of early mobilization through early and intensive occupational therapy (OT), including polysensory stimulation, body positioning, cognitive stimulation exercises, BADLs, upper extremity motor exercises, and family involvement, on non-intubated, elderly patients (≥ 60) in addition to the study center’s standard, non-pharmacological delirium prevention care [21]. Delirium associated outcomes included incidence, duration, and severity; they found significant differences in incidence and duration of delirium, with p-values of 0.001 and <0.001 respectively, but no significant difference in severity (Tables 3-5).

**Table 4.** Results on Duration of Delirium

| **Duration** |  |  |  |  |
| --- | --- | --- | --- | --- |
| **Intervention** | **Number of patients enrolled** | **Duration measurement** | **Group Results** | **Statistical Results** |
| ABCDE bundle [23] | n=296<br>(CG n=146; IG n=150) | duration in days<br>median (IQR) | CG 3 (1, 6)<br>IG 2 (1, 4) | p=0.52 |
|  |  | % ICU days spent delirious<br>median (IQR) | CG 50% (30, 64.3)<br>IG 33.3% (18.8, 50) | p=0.003* |
| ABCDE bundle [24] | | number of days a patient had<br>positive ICDSC score<br>mean ± SD (range) | CG 3.8 ± 2.9 (1.0, 14.0)<br>IG 1.72 ± 0.8 (1.0, 4.0) | p<0.001* |
| Automated reorientation [41] | n=30<br>(CG n=10; UG n=10; FG n=10) | delirium-free days<br>mean (SD) | CG 1.6 (1.13)<br>UG 1.6 (1.07)<br>FG 1.9 (0.99) | p=0.0437* |
|  |  | days of delirium<br>mean (SD) | CG 0.9 (1.28)<br>UG 0.6 (0.84)<br>FG 0.3 (0.48) | p>0.05 |
| Dynamic lighting [45] | n=734<br>(CG n=373; IG n=361) | duration in hours<br>median (IQR) | CG 2 (1, 5)<br>IG 2 (2, 5) | p=0.87 |
| Family, caregiver [39] | n=30<br>(CG n=14; IG n=16) | duration in days<br>mean (SD) | CG 4.14 (4.04)<br>IG 1.94 (1.34) | -- |
| Mirrors [31] | n=223<br>(CG n=108; IG n=115) | duration in days<br>median (IQR [range]) | CG 2 (1, 8 [1, 3])<br>IG 1 (1, 3 [1, 25]) | RR 0.66 (0.26, 1.70) †<br>p=0.393 |
| | | proportion of ICU stay<br>mean (SD) | CG 0.65 (0.29)<br>IG 0.54 (0.30) | Co-eff -0.10 (-0.67, 0.47) †<br>p=0.729 |
| Mobility, early & intensive OT [21] | n=140<br>(CG n=70; IG n=70) | ratio of delirium duration to<br>exposure time | CG IRR 6.66 (5.23, 8.3) †<br>IG IRR 0.15 (0.12, 0.19) † | CG p=0.000*<br>IG p=0.000* |
| Mobility, early mobility protocol [26] | n=58<br>(CG n=27; IG n=31) | duration in days<br>mean ± SD (range) | CG 2.70 ± 2.18 (0, 9)<br>IG 3.58 ± 2.68 (0, 9) | p=0.18 |
| Mobility, FES cycling therapy [42] | n=16<br>(CG n=8; IG n=8) | duration in days<br>median (IQR) | CG 6.0 (3.3, 13.3)<br>IG 0.0 (0.0, 3.0) | p=0.042* |
| Mobility, ROM exercises [35] | n=94<br>(CG n=47; IG n=47) | duration in hours<br>median (range) | CG 38 (9, 120)<br>IG 15 (3, 144) | Z= -0.997 p>0.05 |
| MORE bundle + nursing education [43] | n=483<br>(CG n=230; IG n=253) | duration in hours<br>median (IQR) | CG 20 (9.5, 37) 16.1% of ICU LOS<br>IG 16 (8, 24) 9.6% of ICU LOS | p<0.001* |
| Multicomponent bundle (staff education & environmental changes) [22] | n=148<br>(CG n=69; IG n=79) | % of days with delirium<br>mean ± SD | CG 35.84 ± 39.31<br>IG 26.18 ± 35.38 | $p=0.001^*$ |
| Multicomponent bundle + education [25] | n=123<br>(CG n=57; IG n=66) | delirium-free days out of 30<br>mean (range) | CG 24 (22, 26) †<br>IG 27 (25, 28) † | $p=0.002^*$ |
| Multicomponent non-pharmacologic bundle [32] | n=160<br>(CG n=79; IG n=81) | duration in hours<br>mean ± SD | CG 60.2 ± 15.8<br>IG 28.1 ± 8.6 | $p<0.001^*$ |
| Multicomponent bundle + nursing education [46] | n=89<br>(CG n=38; IG n=51) | number of delirious days<br>mean ± SD | CG 8.2 ± 5.7<br>IG 4.5 ± 4.4 | $p<0.001^*$ |
| Risk factor screening & target modification [47] | n=278<br>(CG n=137; IG n=141) | duration in days, from first positive CAM-ICU to recovery (2 consecutive days with negative CAM-ICU) | all durations (1-5 days), both groups | $p=0.876$ |

**Abbreviations:** CG: control group; FES: functional electrical stimulation; FG: family voice group; IG: intervention group; IQR: interquartile range; IRR: incidence risk ratio; OT: occupational therapy; ROM: range of motion; RR: relative risk; SD: standard deviation; UG: unknown voice group.
**Legend:** \* = significant, $p<0.05$ ; † = 95% confidence interval.

**Table 5.** Results on Severity of Delirium

| **Severity** |  |  |  |  |  |
| --- | --- | --- | --- | --- | --- |
| **Intervention** | **Number of patients enrolled** | **Measurement** | **Group Results** |  | **Statistical Results** |
| Family, caregiver [39] | n=30<br>(CG n=14; IG n=16) | DI<br>mean (SD) | Overall<br>(Days 1-3) | NA | p=0.27 |
|  |  |  | Day 1 | CG 2.07 (4.05)<br>IG 10.56 (3.5) | NA |
|  |  |  | Day 2 | CG 8 (6.34)<br>IG 5.38 (5.45) | NA |
|  |  |  | Day 3 | CG 5.5 (7)<br>IG 3.43 (4.96) | NA |
| Mobility, early & intensive OT [21] | n=140<br>(CG n=70; IG n=70) | DRS<br>mean (range) |  | CG 10 (8, 13) †<br>IG 9 (6, 12) † | p=0.7 |
| Risk factor screening & target modification [47] | n=278<br>(CG n=137; IG n=141) | DRS-R-98<br>number (%) | Overall (Results from all 3 severity groups) | NA | Z= -0.792<br>p=0.428 |
|  |  |  | Mild | CG 10 (7.30)<br>IG 7 (4.96) | NA |
|  |  |  | Moderate | CG 21 (15.33)<br>IG 8 (5.67) | NA |
|  |  |  | Severe | CG 10 (7.30)<br>IG 4 (2.84) | NA |
| Roy adaptation model [33] | n=100<br>(CG n=50; IG n=50) | NEECHAM<br>Confusion Scale | Day 1-3, AM & PM<br>(6 time points) | See full reference text. | p>0.05 |
|  |  |  | Day 4-7, AM & PM<br>(8 time points) | See full reference text. | p≤0.028*<br>(range p=0.000 to p=0.028) |

**Abbreviations:** DI: Delirium Index; DRS: Delirium Rating Scale; DRS-R-98: Delirium Rating Scale-Revised-98; NA: not analyzed; OT: occupational therapy.
**Legend:** \* = significant difference, p<0.05; † = confidence interval not specified.

The study by Karadas and Ozdemir assessed the effect of range of motion exercises on delirium in elderly patients (≥65 years) [35]. Interventional care included range of motion exercises for 30 minutes daily after establishing the patient’s ability to complete 10 repetitions on each of the four extremities while lying in bed. They reported no statistically significant differences between cohorts for delirium associated outcomes (Tables 3 & 4).

Campbell addressed early mobilization in mechanically ventilated ICU patients [26]. They measured the effect of a tiered protocol of ROM exercises, bed mobility exercises, seated balance activities, transfer activities (such as bed to chair), standing exercises, and ambulation on delirium incidence and duration but found neither to be significant (Tables 3 & 4).

The effectiveness of functional electrical stimulation (FES) to promote mobility and recovery in patients with sepsis was evaluated by Parry et al. [42]. The intervention included use of a motorized cycle ergometer to directly stimulate four major lower limb muscles (quadriceps, hamstrings, gluteals, and calves) five times weekly for 20-60 minutes a session dependent on the individual patient’s tolerance. While delirium incidence was not significantly affected (Table 3), the median days of delirium differed between arms (6.0 in control and 0.0 in intervention) (Table 4).

##### 3.2.2 Family involvement

Of the 12 studies in our review which focused on individual interventions, three studied the effect of family involvement on delirium in adult ICU patients. One was a randomized pilot study [39], one a RCT [29], and one was a pre-post study [44]. All three studies utilized CAM-ICU in their assessment of delirium.

Mailhot et al. constructed a randomized pilot study to explore the effect of a FC assisting with delirium management in the MENTOR_D intervention, measuring the outcomes of duration, occurrence, and severity over three days [39]. This intervention enrolled 14 control dyads and 16 patient-FC dyads, which had the FC apply bedside strategies to aid the patient in reorientation. In addition to reorientation, the FC was asked to observe and communicate signs of delirium with nursing staff, present family memories, and speak clearly and simply. Delirium duration and occurrence on post-operative Day 2 improved clinically between groups (duration, mean days from 4.14 to 1.94; occurrence, from 71.40% to 43.80%); however, this result was not assessed for statistical significance and the severity result was not found to be significant (Tables 3-5).

The RCT performed by Eghbali-Babadi et al. investigated a modified family visitation policy, implementing an additional 30-40 minute special visit by an approved family member, and its effect on delirium incidence [29]; they found a statistically significant reduction in delirium incidence in the intervention group with a p-values of 0.04, <0.05, and 0.03 at three different time points (Table 3).

Rosa et al. also measured the effect of a modified family visitation policy on delirium incidence [44]. Their pre-post study included the extension of visitation hours from 4.5 hours per day over three visitation blocks to 12 hours per day between 09:00-21:00. This resulted in a statistically significant difference in delirium incidence, improving from 20.5% to 9.6% (Table 3).

#### 3.2.3 Environmental (lighting, music therapy, automated reorientation)

Three RCTs assessed the impact of environmental factors on delirium in the ICU, assessed by CAM-ICU, through manipulation of light [45], music therapy [28], or automated reorientation [41].

In Simons et al.’s dynamic lighting application (DLA) trial, patients were exposed to bright, high intensity lighting while delirium incidence and duration were measured [45]. The intervention group was exposed to a peak of 1700 lux/4300 K from 09:00-11:30 and 13:30-16:00, and a daytime minimum of 300 lux/3000 K from 11:30-13:30; the control group was exposed to 300 lux/3000 K (Table 1). Neither the cumulative incidence of ICU-acquired delirium nor the duration were significantly affected, and the trial was ended early after the intervention was deemed futile (Tables 3 & 4).

Damshens et al. introduced therapeutic music, selected by a music expert, twice a day for 45 minutes to assess the effect on delirium incidence in the ICU [28]. Patients in the control group received conventional care for the duration of their admission. Delirium incidence did not demonstrate any changes between the two groups (Table 3).

Munro et al. developed a new patient reorientation strategy which utilized bilingual (Spanish or English) pre-recorded messages by either family members or females unknown to the subjects [41]. The recordings included an introduction with the patient’s name and location, and several additional randomly ordered statements were included in order to reorient the patient to their unfamiliar surroundings and reason for hospitalization. All three arms (2 intervention groups and 1 control group) were compared and it was found that the family voice group had a significant improvement in delirium free days (p= 0.0437) but not mean days of delirium (Table 4).

#### 3.2.4 Self-involvement approaches (mirror usage, mindfulness exercises)

The remaining two studies on the effect of individual interventions assessed the impact of self-involvement approaches, including mirror usage [31] and mindfulness exercises [38], on ICU delirium measured by CAM-ICU. One study was a pilot RCT [31], while the other was a mixed-methods pilot study [38].

Giraud et al. tested the effect of introducing structured mirror usage into post-operative recovery in elderly ICU patients (≥70 years) after cardiac surgery [31]. Mirror usage was standardized by developing a protocol for nurses and physiotherapists, aiming to use both small, personal mirrors as well as larger posture mirrors in order to help the patient with reorientation and self-awareness, enhance multisensory feedback on minor procedures, and augment passive and active physical therapies. The control cohort received usual care, including allowing control patients who brought a mirror from home to use it per their normal habits. After comparing the usual care group with the mirrors group, no significant improvement was found in delirium incidence, ICU days with delirium, or proportion of ICU length of stay with delirium (Tables 3 & 4).

The study by Lisann-Goldman et al. had subjects 40 years or older participate in Langerian mindfulness discussion exercises both prior to and after elective cardiac surgery with cardiopulmonary bypass [38]. In addition to discussion exercises, patients listened to an audio file before surgery. This audio file walked them through techniques on how to re-assess one’s situation and outlook in terms of self-related-to-environment, encouraging the patients to focus on the process of change and allowing oneself to accept new ideas and take control over the unknown. The discussion exercises continued post-operatively twice daily. In contrast, the ‘informational control’ group went through normal pre-operative discussions followed by an audio file describing the process of cardiac surgery. They found that no subject developed delirium in either the interventional or the ‘informational control’ group so the effectiveness of the treatment could not be assessed.

### 3.3 Bundled protocols

#### 3.3.1 ‘Wake Up and Breathe’ protocol

Khan et al. designed a ‘Wake Up and Breathe’ protocol in a pre-post interventional study to assess for any change in delirium and sedation in mechanically ventilated, adult ICU patients [36]. They modified elements of the ABC to implement a spontaneous awakening trial and daily sedation vacation followed by a spontaneous breathing trial depending on the patient’s response [48]. Delirium was assessed by CAM-ICU, and both incidence and prevalence of delirium were analyzed, finding no significant change in either measured outcome (Table 3).

#### 3.3.2 ABCDE/ABCDEF bundles

Five of the 15 studies which examined delirium bundles studied the effectiveness of ABCDE or ABCDEF bundle protocols on reducing delirium. ABCDE/F bundles had multiple components which included spontaneous awakening (A) and breathing (B) trials, interdisciplinary coordination of sedatives and medications (C), delirium screening and management (D), early mobilization (E), and family engagement and involvement (F). Of these five studies, two were pre-post studies [23, 37], one was a prospective cohort study [34], one was a quasi-experimental quality improvement project [27], and one was a retrospective cohort study [24]. Three measured delirium outcomes using CAM-ICU [23, 27, 37] and two utilized ICDSC [24, 34].

Balas et al. assessed the impact of an ABCDE bundle on in adult ICU patients evaluating the prevalence and duration of delirium, in both total days and percent of ICU days spent delirious [23]. The prevalence and percent of ICU days spent delirious were improved in the post period with p-values of 0.03 and 0.003 respectively (Table 3 & 4). However, the overall duration of delirium was not significantly different (Table 4).

The retrospective assessment of an ABCDE bundle by Bounds et al. assessed its effect on delirium prevalence and duration in an adult ICU population [24]. Both the prevalence and duration were significantly decreased in the ABCDE bundle group (p= 0.01 and 0.001 respectively; Tables 3 & 4).

Kram et al. also looked at a similar patient cohort, all adult patients 18 or older admitted to the ICU, in a pre-post ABCDE bundle study although with a smaller subject population (n=83; Balas, n=296; Bounds, n=159) [37]. They assessed the effectiveness of the ABCDE bundle on delirium by measuring delirium prevalence and comparing it to a control based on literature values. The measured delirium prevalence of 19% (Table 3) fell outside the cited literature values of 20-80%.

The prospective cohort study by Hsieh et al. implemented the ABCDE bundle in stages, as follows: the baseline cohort (B) already had the B component established in critical care; the B-AD cohort had the AD component introduced in a second phase of implementation; and the B-AD-EC cohort had the full bundle as part of their routine critical care [34]. Despite a thorough study and the large number of measured outcomes, the only delirium-specific outcome they assessed was percent of patients ever recorded with a positive CAM-ICU result (Table 3). This was not analyzed for significance, but the incidence rates did improve across groups (B: 71%, B-AD: 48%, and B-AD-EC: 45%).

Chai initiated an ABCDEF bundle in a mixed ICU setting and analyzed delirium incidence in the adult patients [27]. Delirium incidence was compared between morning and night occurrences (morning 08:00-11:00; night 20:00-23:00); both showed significant improvement in the intervention group with a p-value <0.001 for both morning and evening measurements (Table 3).

#### 3.3.3 Other bundled protocols

The remaining nine bundle studies developed new, unique bundles. They included four pre-post studies [25, 43, 46, 47], three RCTs [32, 33, 40], one quasi-experimental study [22], and one action research study [30]. Seven assessed delirium incidence and duration using CAM-ICU [22, 25, 30, 32, 40, 46, 47], one used NEECHAM [33], and one used ICDSC [43].

A quasi-experimental study designed by Arbabi et al. developed a multi-component delirium management bundle comprised of staff education and environmental and non-pharmacologic care changes [22]. They measured the effectiveness of their bundle by comparing delirium incidence and duration for both groups, finding a significant difference in both outcomes (p = 0.01 and 0.001 respectively; Table 3 & 4).

Bryczkowski et al. assessed the effectiveness of their bundle, which included a staff-patient-family education program, medication management strategies, and non-pharmacological sleep enhancement protocols, on delirium incidence and delirium free days in patients over the age of 50 years [25]. The research team found no significant improvement in delirium incidence (Table 3), although the average total number of delirium-free days out of 30 changed significantly from 24 to 27 between groups (p=0.002; Table 4).

Another bundle study developed by Fallahpoor et al. focused specifically on adults admitted to the ICU after elective CABG. This post-CABG delirium management bundle was assessed in an action research study and had three elements focusing on pre-, intra-, and post-operative methods to identify delirium risk factors, optimize time spent in surgery, and introduce staff education and post-operative environmental changes [30]. Delirium related outcomes included incidence ratio and total number of recorded delirium events, with significant differences found in both (p=0.001 and 0.008; Table 3).

In the RCT conducted by Guo et al. the effect of a bundle consisting of cognitive prehabilitation, post-operative cognitive stimulation activities, environmental changes, music therapy, and non-pharmacologic care changes on delirium incidence and duration after oral tumor resection was studied [32]. The incidence of delirium improved significantly overall, but was only significantly different on post-operative Day 1 compared to Days 2 and 3 (p=0.035, p=0.374, p=0.364 respectively; Table 3); the duration of delirium also differed significantly (p< 0.001; Table 4).

Hamzehpour et al. designed an RCT and implemented the Roy adaptation nursing model, which focuses on balance of nutrition, electrolytes, and fluids while promoting activity, sleep hygiene, and monitoring of circulation and endocrine function, into their critical care protocol [33]. Their primary delirium-specific outcomes were incidence and severity, and they analyzed both outcomes for two time points (morning & night) for seven days. Their research only showed significant improvements to incidence on Day 7, both morning and night (p<0.008 and p<0.05; Table 3), but delirium severity, assessed with NEECHAM, improved through the morning of Day 4 to the night of Day 7 at all measured time points (every time point, p≤0.028; Table 5).

Moon and Lee implemented a bundle which included early cognitive assessments and reorientation, sensory aids, environmental changes, consistent care staff and location, familiar items from home, nursing care changes, and early mobility as part of an RCT aimed at assessing differences in delirium incidence [40]. Their study did not show a significant difference between the intervention and the control group (Table 3).

In a pre-post, observational quality improvement project, Rivosecchi et al. combined staff education with a non-pharmacologic bundle to look at incidence and duration of delirium in any adult patient aged 18 or older admitted to the medical ICU [43]. Their M.O.R.E. bundle included (M)usic, (O)pening blinds, (R)eorientation and cognitive stimulation, and (E)ye and ear care. Both incidence and duration were significantly impacted, with incidence decreasing from 15.7% to 9.4% and a reduction in duration from 16.1% of the ICU stay to 9.6% (p = 0.04 and <0.001 respectively; Tables 3 & 4).

Sullinger et al. enrolled critically ill patients with acute delirium in a pre-post retrospective study, tailoring their bundle to incorporate staff education with sensory aids, healing arts techniques, mobility, environmental changes, and family presence [46]. Their bundle also included the initiation of anti-psychotic medications if non-pharmacologic tactics failed. The only specifically delirium related outcome was the number of days spent delirious, resulting in a significant decrease from 8.2 to 4.5 median days (Table 4).

Any patient 18 years or older admitted to a cardiothoracic ICU after CABG surgery was analyzed for incidence, duration, and severity of delirium by Zhang et al. in a prospective pre-post study [47]. Their delirium bundle targeted risk factor screening and modifications, including increased family visits, reorientation, and changes to nursing care. The only significant improvement was to incidence of delirium which dropped from 29.93% to 13.48% (Table 3), while the intervention had no impact on duration or severity (Tables 4 & 5).

## 4. Discussion

### 4.1 Summary of Findings

Our review included 27 trials that evaluated the effect of various non-pharmacological treatment and management protocols on delirium in an ICU setting. Assessment of the efficacy of these protocols in the last five years was most commonly done by considering incidence and/or prevalence. Twenty-five studies assessed for the effects on incidence and/or prevalence, with eleven studying individual approaches and 14 studying bundles. Of these 25 trials, eleven found significant improvement overall [21-24, 27, 29, 30, 34, 43, 44, 47], nine found no significant improvement [25, 26, 28, 31, 35, 36, 40, 42, 45], and two only found significant change at certain time points [32, 33]; the remaining three did not analyze for statistical significance of their results [37-39]. The eleven effective interventions for incidence and/or prevalence were primarily bundled protocols (8 trials) [22-24, 27, 30, 34, 43, 47], followed by family approaches (2 trials) [29, 44], and early and intensive OT (1 trial) [21]. The two studies with time point dependent changes were both bundles, one non-pharmacologic [32]and the other introduced the Roy adaptation nursing model[33]. The multicomponent non-pharmacologic bundle found improvements in incidence both overall and on Day 1, while the Roy adaptation trial only saw a change in incidence on Day 7 in both the morning and the evening. Three studies did not analyze for statistical significance [37-39]; however, the family caregiver intervention saw an overall reduction in the percent of subjects who developed delirium from 71.40% to 43.80% [39]. The study on mindfulness exercises had no subjects in either investigational group develop delirium [38], and an ABCDE bundle reported a post-bundle incidence of 19% but stated there was no pre-bundle data with which to compare [37]. No other studies that looked at delirium incidence were effective.

In addition to incidence and prevalence, another common outcome was duration of delirium. Sixteen of the reviewed studies evaluated duration of delirium, eight focusing on individual interventions and eight introducing bundled protocols. Of these 16 studies, eight found significant changes overall [21, 22, 24, 25, 32, 42, 43, 46], two had significant improvements at select time points [23, 41], and five did not have significant results [26, 31, 35, 45, 47]; the one remaining study did not assess for statistical significance [39]. Six of the eight successful trials were bundles and both of the two effective individual therapies were mobility-focused (early and intensive OT, and FES) [21, 42]. Only four studies looked at delirium severity [21, 33, 39, 47] with only one finding any significant results, and only finding them at select time-points (Roy Adaptation Model) [33].

The pilot RCT performed by Alvarez et al. utilized a unique method for assessing the performance of their intervention. In addition to assessing delirium incidence, they measured the ratio of delirium duration to the amount of time exposed to the treatment (IRR) [21]. They found that IRR decreased as the time exposed to treatment increased to a significant degree (p= 0.000). This ratio could be explained in three ways. Either the duration of delirium stayed the same as the time exposed to treatment increased, the duration of delirium increased slower than the time exposed increased, or the duration of delirium decreased while the time to exposure increased. However, the last explanation is impossible, due to the duration of delirium being a sum overtime which could not decrease, such that the result must be explained by either a small increase or no increase in the duration of delirium. If the decrease in the IRR is explained by smaller and smaller increases to delirium duration it is likely that the IRR results from either the trend of patients slowly becoming healthier over time, or the conjunction of that with the intervention. However, if the IRR is explained by delirium duration ceasing to increase, then once it stops the treatment may still be effective but is no longer becoming more effective over time and plateaus in effectiveness.

#### 4.2 Implications of Results and Application to Practice

Studies focused on individual interventions had a wide range of limitations and were, on the whole, less effective than bundled protocols in the treatment and management of delirium. Many of these studies had limited reliability due to small or extremely small sample sizes [26, 38, 41, 42]. Additionally, even when results were significant, they often had a limited application to practice due to the prevalence of restricted populations. Three of the individual intervention studies limited their study cohort to elderly adults [13, 21, 35], a population which is at an increased risk for delirium. It is unclear whether these results would apply to younger patients. Studied populations were also commonly narrowed to either exclusively intubated [42] or non-intubated patients [21, 29] or patients with a particular illness [31, 39, 42]. Another possible limitation was a questionable reliability of delirium assessment. This is mentioned by Campbell who stated that 35% of CAM-ICU were incorrectly labeled as ‘unable to assess’ [26]. The question of reliability was also raised in Lisann-Goldman et al.. This study could not assess the effectiveness of their intervention due to no patients developing delirium [38]. However, this could be explained by the fact that fully sedated patients were considered as ‘not delirious’ since CAM-ICU could not be performed. The authors also noted that since it often takes weeks or months to fully integrate new behavioral thought techniques, that a study focused on changing thought patterns in days would not entirely reflect the full benefit if any were present [38].

Eight of the individual intervention studies were RCTs. This type of study introduces the possibility of additional limitations due to the nature of its design. Two of these RCTs had a high risk of bias [29, 41] due to failure to blind patients and personnel as well as blinding of the outcome assessment and improper allocation concealment. The question of blinding raises another possible limitation to many of these studies, namely the possibility of the Hawthorne effect in patients who knew that they were being observed and receiving an intervention for the treatment/ prevention of delirium.

The fifteen studies which investigated bundled protocols had, overall, larger sample sizes, fewer cohorts with limited populations, and better reliability of delirium assessment than the studies which focused on individual interventions. The smallest sample size was 83 patients [37]; however, this study did not split their sample into multiple cohorts and all patients received the intervention. One study had a sample size of 89 [46], and all other studies had a sample size of at least 100 patients. A total of five studies restricted their studied population beyond adult ICU patients [25, 30, 32, 36, 47]. Two of these limited their population by age [25, 32]; however, Bryczkowski et al., despite limiting their population by age, included any patients greater than 50 years old, younger than the age when delirium risk is noted to increase [49, 50]. One study limited its population to mechanically ventilated patients [36], two considered only patients undergoing CABG[30, 47], and one studied patients after oral tumor resection[32]. These population restrictions could limit the generalizability and applicability of the interventions; however, this risk is reduced since bundles were often investigated in multiple studies with similar results.

Only three of the bundle studies were RCTs [32, 33, 40]. Each of these RCTs had an unclear risk of bias with the most common risk being the inability to blind participants and personnel. The impossibility of blinding in delirium intervention studies makes RCTs a questionable approach. Eight of the bundle studies, recognizing blinding as an impossibility, chose to conduct pre-post prospective studies rather than RCTs [23, 25, 27, 36, 37, 43, 46, 47]. These studies carried a lower risk of introducing bias to their studies and avoided crossover between arms. The pre-post study performed by Kram et al. had the major limitation of not including a pre cohort and only comparing the results of their intervention with literature values [37]. Additionally, while they found their measured delirium prevalence to fall outside their included literature values (19%), this prevalence falls within the values provided by the current literature (19-87%) [2]. Chai’s pre-post study had questionable reliability of delirium assessment. While all other studies assessed delirium whenever RASS was ≥ −3, they reported that patients were unable to be assessed whenever RASS was < −2, resulting in a greater proportion of patients not assessed for delirium [27].

Given the findings of this systematic review, further research is warranted in order to confirm these results and apply them to other patient populations. Multicomponent, bundled approaches were more successful at improving delirium outcomes compared to individual techniques; however, the effective individual tactic of family engagement was included as a component in the effective bundles. Although a majority of the reviewed bundles were effective, it is difficult to compare results as the trials had large differences in study design, enrollment numbers, and delirium assessment measures.

### 4.3 Strengths and Limitations

The strengths of our systematic review include thorough search terms and methodology to assess a vast majority of recent literature in this field.

One limitation of this systematic review is that we only focused on trials within the past five years which excluded some well-cited early studies on delirium. We also did not evaluate other listed outcomes which could provide additional insight into any change in delirium status. Since the condition can be transient and delirium screenings are not performed as frequently throughout the ICU day as other measurements, outcomes such as restraint use or amount of prescribed sedatives or anti-psychotic medications would be beneficial to assess in this setting. While the decision to omit exclusion criteria on study design allowed for assessment of a broader range of trials, it was difficult to compare outcomes when multiple differing designs and measurement tools were used. Although the CAM-ICU was widely used, some studies used alternative tools and there was no standardized way of defining or measuring delirium duration or severity. A different measurement tool was used to evaluate severity in each of the four studies reviewing this outcome, and duration was defined in a multitude of fashions. Combining this realization with the fact that some studies focused on highly specific subpopulations suggests that some trials may need to be replicated in a standardized fashion to account for any differences in methodology or subjective assessments.

## 5. Conclusions

Many ICU delirium treatment and management protocols were developed and tested within the last five years in a variety of study designs. Few trials on individual interventions had positive effects on delirium incidence and duration, but multicomponent bundles were found to be more effective overall while incorporating the effective individual intervention of family engagement. Based on the results of bundle studies, the implementation of multicomponent protocols in ICUs can reduce ICU delirium, thereby reducing cost of care, improving overall outcomes, and limiting time spent mechanically ventilated, medicated, or admitted. Despite these results, further research is needed on individual interventions in order to improve specific elements of multicomponent bundles by adding or removing ineffective therapies. Additional research is also warranted to evaluate for any positive effects in more generalized hospital populations.

## Data Availability

NA

**Abbreviations**
BADLs: basic activities of daily living;
CABG: coronary artery bypass graft;
CAM: Confusion Assessment Method;
DI: Delirium Index;
DLA: dynamic lighting application;
DRS: Delirium Rating Scale;
DRS-R-98: Revised Delirium Rating Scale;
FES: functional electrical stimulation;
ICDSC: Intensive Care Delirium Screening Checklist;
ICU: intensive care unit;
NEECHAM: NEElon and CHAMpagne;
OT: occupational therapy;
PRISMA: Preferred Reporting Items for Systematic Reviews and Meta-Analyses;
RCT: randomized controlled trial;
ROM: range of motion.

---

Supplementary Appendix 1 - PRISMA Checklist

Supplementary Appendix 2 - Database Search Terms

Supplementary Table 1 - Data Extraction Form

Supplementary Table 2 - Risk of Bias Assessment Form

## References

1. W.-L. Lin, Y.-F. Chen and J. Wang, Factors associated with the development of delirium in elderly patients in intensive care units, Journal of Nursing Research. 4 (2015) 322–329.

2. J. Kalabalik, L. Brunetti and R. El-Srougy, Intensive care unit delirium: a review of the literature, Journal of pharmacy practice. 2 (2014) 195–207.

3. V. J. Page, E. W. Ely, S. Gates, et al., Effect of intravenous haloperidol on the duration of delirium and coma in critically ill patients (Hope-ICU): a randomised, double-blind, placebo-controlled trial, The Lancet Respiratory Medicine. 7 (2013) 515–523. https://doi.org/10.1016/S2213-2600(13)70166-8

4. A. M. Parker, T. Sricharoenchai, S. Raparla, et al., Posttraumatic Stress Disorder in Critical Illness Survivors: A Metaanalysis*, Critical Care Medicine. 5 (2015) 1121–1129. 10.1097/ccm.0000000000000882

5. I. Lat, W. McMillian, S. Taylor, et al., The impact of delirium on clinical outcomes in mechanically ventilated surgical and trauma patients, Critical care medicine. 6 (2009) 1898–1905.

6. J. R. Maldonado, Delirium pathophysiology: An updated hypothesis of the etiology of acute brain failure, International Journal of Geriatric Psychiatry. 11 (2018) 1428–1457. 10.1002/gps.4823

7. S. T. Micek, N. J. Anand, B. R. Laible, et al., Delirium as detected by the CAM-ICU predicts restraint use among mechanically ventilated medical patients*, Critical Care Medicine. 6 (2005) 1260–1265. 10.1097/01.ccm.0000164540.58515.bf

8. S. Ahmed, B. Leurent and E. L. Sampson, Risk factors for incident delirium among older people in acute hospital medical units: a systematic review and meta-analysis, Age and Ageing. 3 (2014) 326–333. 10.1093/ageing/afu022

9. T. G. Fong, S. R. Tulebaev and S. K. Inouye, Delirium in elderly adults: diagnosis, prevention and treatment, Nature Reviews Neurology. 4 (2009) 210–220. 10.1038/nrneurol.2009.24

10. B. Van Rompaey, M. M. Elseviers, M. J. Schuurmans, et al., Risk factors for delirium in intensive care patients: a prospective cohort study, Critical Care. 3 (2009) R77. 10.1186/cc7892

11. R. P. Thom, M. P. Bui, B. Rosner, et al., A Comparison of Early, Late, and No Treatment of Intensive Care Unit Delirium With Antipsychotics: A Retrospective Cohort Study, Prim Care Companion CNS Disord. 6 (2018) 10.4088/PCC.18m02320

12. M. P. Knauert and M. A. Pisani, Dexmedetomidine for hyperactive delirium: worth further study, J Thorac Dis. 9 (2016) E999-E1002. 10.21037/jtd.2016.08.14

13. T. D. Girard, M. C. Exline, S. S. Carson, et al., Haloperidol and Ziprasidone for Treatment of Delirium in Critical Illness, N Engl J Med. 26 (2018) 2506–2516. 10.1056/NEJMoa1808217

14. S. Chen, L. Shi, F. Liang, et al., Exogenous Melatonin for Delirium Prevention: a Meta-analysis of Randomized Controlled Trials, Mol Neurobiol. 6 (2016) 4046–4053. 10.1007/s12035-015-9350-8

15. R. C. Arora and C. Cunningham, Losing Sleep Over Delirium, Crit Care Med. 6 (2018) 1036–1038. 10.1097/CCM.0000000000003141

16. D. Moher, A. Liberati, J. Tetzlaff, et al., Preferred reporting items for systematic reviews and meta-analyses: the PRISMA statement, Annals of internal medicine. 4 (2009) 264–269.

17. T. C. Collaboration, Data Colllection Form, (2011)

18. J. P. Higgins, Sterne, J. AC., Savovic, J., Page, M. J., Hróbjartsson, A., Boutron, I.,… Eldridge, S., A revised tool for asessing risk of bias in randomized trials, Cochrane Database of Systemaic Reviews. Suppl 1 (2016) 29–31.

19. S. J. Schaller, M. Anstey, M. Blobner, et al., Early, goal-directed mobilisation in the surgical intensive care unit: a randomised controlled trial, The Lancet. 10052 (2016) 1377–1388. https://doi.org/10.1016/S0140-6736(16)31637-3

20. J. Patel, J. Baldwin, P. Bunting, et al., The effect of a multicomponent multidisciplinary bundle of interventions on sleep and delirium in medical and surgical intensive care patients, Anaesthesia. 6 (2014) 540–549. 10.1111/anae.12638

21. E. A. Álvarez, M. A. Garrido, E. A. Tobar, et al., Occupational therapy for delirium management in elderly patients without mechanical ventilation in an intensive care unit: A pilot randomized clinical trial, Journal of Critical Care. (2017) 85–90. https://doi.org/10.1016/iicrc.2016.09.002

22. M. Arbabi, J. Zebardast, A. A. Noorbala, et al., Efficacy of Liaison Education and Environmental Changes on Delirium Incidence in ICU, Arch Neurosci. 2 (2018) e56019. 10.5812/archneurosci.56019

23. M. C. Balas, E. E. Vasilevskis, K. M. Olsen, et al., Effectiveness and safety of the awakening and breathing coordination, delirium monitoring/management, and early exercise/mobility bundle, Critical care medicine. 5 (2014) 1024–1036. 10.1097/CCM.0000000000000129

24. M. Bounds, S. Kram, K. G. Speroni, et al., Effect of ABCDE Bundle Implementation on Prevalence of Delirium in Intensive Care Unit Patients, Am J Crit Care. 6 (2016) 535–544. 10.4037/ajcc2016209

25. S. B. Bryczkowski, M. C. Lopreiato, P. P. Yonclas, et al., Delirium prevention program in the surgical intensive care unit improved the outcomes of older adults, Journal of Surgical Research. 1 (2014) 280–288. https://doi.org/10.1016/Mss.2014.02.044

26. M. Campbell, 728: THE EFFECT OF AN EARLY MOBILITY PROTOCOL IN CRITICALLY ILL, MECHANICALLY VENTILATED PATIENTS, Critical Care Medicine. 12 (2014) A1535. 10.1097/01.ccm.0000458225.10153.e3

27. J. Chai, The effect of the ABCDEF bundle on incidence of delirium in critically ill patients, ProQuest Dissertations Publishing. (2017) 10286721

28. M. Damshens, M. Sanie, S. Javadpour, et al., The Role of Musicon the Delirium in Traumatic Patients: A Case Study in the ICU of Peymanieh Hospital of Jahrom, Fars Province, Iran, Ambient Science. (2018) 10.21276/ambi.2018.05.sp1.ra11

29. M. Eghbali-Babadi, N. Shokrollahi and T. Mehrabi, Effect of Family-Patient Communication on the Incidence of Delirium in Hospitalized Patients in Cardiovascular Surgery ICU, Iran J Nurs Midwifery Res. 4 (2017) 327–331. 10.4103/1735-9066.212985

30. S. Fallahpoor, H. Abedi and M. Mansouri, Development and Evaluation of Care Programs for the Delirium Management in Patients after Coronary Artery Bypass Graft Surgery (CABG) (2016)

31. K. Giraud, M. Pontin, L. D. Sharples, et al., Use of a Structured Mirrors Intervention Does Not Reduce Delirium Incidence But May Improve Factual Memory Encoding in Cardiac Surgical ICU Patients Aged Over 70 Years: A Pilot Time-Cluster Randomized Controlled Trial, Front Aging Neurosci. (2016) 228. 10.3389/fnagi.2016.00228

32. Y. Guo, L. Sun, L. Li, et al., Impact of multicomponent, nonpharmacologic interventions on perioperative cortisol and melatonin levels and postoperative delirium in elderly oral cancer patients, Arch Gerontol Geriatr. (2016) 112–7. 10.1016/j.archger.2015.10.009

33. H. Hamzehpour, S. Valiee, D. Roshani, et al., The Effect of Care Plan Based on Roy Adaptation Model on the Incidence and Severity of Delirium in Intensive Care Unit Patients: A Randomised Controlled Trial, Journal of Clinical and Diagnostic Research. (2018) 21. 10.7860/JCDR/2018/36366.12256

34. S. J. Hsieh, O. Otusanya, H. B. Gershengorn, et al., Staged Implementation of Awakening and Breathing, Coordination, Delirium Monitoring and Management, and Early Mobilization Bundle Improves Patient Outcomes and Reduces Hospital Costs, Crit Care Med. 7 (2019) 885–893. 10.1097/ccm.0000000000003765

35. C. Karadas and L. Ozdemir, The effect of range of motion exercises on delirium prevention among patients aged 65 and over in intensive care units, Geriatric Nursing. 3 (2016) 180–185. https://doi.org/10.1016/j.gerinurse.2015.12.003

36. B. A. Khan, W. F. Fadel, J. L. Tricker, et al., Effectiveness of implementing a wake up and breathe program on sedation and delirium in the ICU, Crit Care Med. 12 (2014) e791–5. 10.1097/ccm.0000000000000660

37. S. L. Kram, M. C. DiBartolo, K. Hinderer, et al., Implementation of the ABCDE Bundle to Improve Patient Outcomes in the Intensive Care Unit in a Rural Community Hospital, Dimens Crit Care Nurs. 5 (2015) 250–8. 10.1097/dcc.0000000000000129

38. L. R. Lisann-Goldman, F. Pagnini, S. G. Deiner, et al., Reducing Delirium and Improving Patient Satisfaction With a Perioperative Mindfulness Intervention: A Mixed-Methods Pilot Study, Holistic Nursing Practice. 3 (2019) 163–176. 10.1097/hnp.0000000000000321

39. T. Mailhot, S. Cossette, J. Cote, et al., A post cardiac surgery intervention to manage delirium involving families: a randomized pilot study, Nurs Crit Care. 4 (2017) 221–228. 10.1111/nicc.12288

40. K.-J. Moon and S.-M. Lee, The effects of a tailored intensive care unit delirium prevention protocol: A randomized controlled trial, International Journal of Nursing Studies. 9 (2015) 1423–1432. https://doi.org/10.1016/iiinurstu.2015.04.021

41. C. L. Munro, P. Cairns, M. Ji, et al., Delirium prevention in critically ill adults through an automated reorientation intervention - A pilot randomized controlled trial, Heart & Lung. 4 (2017) 234–238. https://doi.org/10.1016/jhrtlng.2017.05.002

42. S. M. Parry, S. Berney, S. Warrillow, et al., Functional electrical stimulation with cycling in the critically ill: A pilot case-matched control study, Journal of Critical Care. 4 (2014) 695.e1–695.e7. https://doi.org/10.1016/i.icrc.2014.03.017

43. R. M. Rivosecchi, S. L. Kane-Gill, S. Svec, et al., The implementation of a nonpharmacologic protocol to prevent intensive care delirium, J Crit Care. 1 (2016) 206–11. 10.1016/jjcrc.2015.09.031

44. R. G. Rosa, T. F. Tonietto, D. B. da Silva, et al., Effectiveness and Safety of an Extended ICU Visitation Model for Delirium Prevention: A Before and After Study, Crit Care Med. 10 (2017) 1660–1667. 10.1097/ccm.0000000000002588

45. K. S. Simons, R. J. Laheij, M. van den Boogaard, et al., Dynamic light application therapy to reduce the incidence and duration of delirium in intensive-care patients: a randomised controlled trial, Lancet Respir Med. 3 (2016) 194–202. 10.1016/s2213-2600(16)00025-4

46. D. Sullinger, A. Gilmer, L. Jurado, et al., Development, Implementation, and Outcomes of a Delirium Protocol in the Surgical Trauma Intensive Care Unit, Ann Pharmacother. 1 (2017) 5–12. 10.1177/1060028016668627

47. W. Zhang, Y. Sun, Y. Liu, et al., A nursing protocol targeting risk factors for reducing postoperative delirium in patients following coronary artery bypass grafting: Results of a prospective before-after study, International Journal of Nursing Sciences. (2017) 10.1016/j.ijnss.2017.02.002

48. T. D. Girard, J. P. Kress, B. D. Fuchs, et al., Efficacy and safety of a paired sedation and ventilator weaning protocol for mechanically ventilated patients in intensive care (Awakening and Breathing Controlled trial): a randomised controlled trial, Lancet. 9607 (2008) 126–34. 10.1016/S0140-6736(08)60105-1

49. M. Y. Kim, U. J. Park, H. T. Kim, et al., DELirium Prediction Based on Hospital Information (Delphi) in General Surgery Patients, Medicine (Baltimore). 12 (2016) e3072. 10.1097/MD.0000000000003072

50. M. van den Boogaard, P. Pickkers, A. J. Slooter, et al., Development and validation of PRE-DELIRIC (PREdiction of DELIRium in ICu patients) delirium prediction model for intensive care patients: observational multicentre study, BMJ. (2012) e420. 10.1136/bmj.e420

51. J. Barr and P. P. Pandharipande, The Pain, Agitation, and Delirium Care Bundle: Synergistic Benefits of Implementing the 2013 Pain, Agitation, and Delirium Guidelines in an Integrated and Interdisciplinary Fashion, Critical Care Medicine. 9 (2013) S99-S115. 10.1097/CCM.0b013e3182a16ff0

52. R. Cavallazzi, M. Saad and P. E. Marik, Delirium in the ICU: an overview, Annals of intensive care. 1 (2012) 49.

53. E. B. Milbrandt, S. Deppen, P. L. Harrison, et al., Costs associated with delirium in mechanically ventilated patients, Critical care medicine. 4 (2004) 955–962.

54. A. Morandi, N. E. Brummel and E. W. Ely, Sedation, delirium and mechanical ventilation: the ‘ABCDE’ approach, Current Opinion in Critical Care. 1 (2011) 43–49. 10.1097/MCC.0b013e3283427243

55. B. T. Pun, M. C. Balas, M. A. Barnes-Daly, et al., Caring for Critically Ill Patients with the ABCDEF Bundle: Results of the ICU Liberation Collaborative in Over 15,000 Adults, Critical Care Medicine. 1 (2019) 3–14. 10.1097/ccm.0000000000003482

56. X. Huang, J. Lin, & D. Demner-Fushman, Evaluation of PICO as a knowledge representation for clinical questions, AMIA… Annual Symposium proceedings. (2006) 359–363.

